# InSleep46: Deployment of a remote monitoring device for the detection and monitoring of dementia risk in older adult populations: a feasibility study

**DOI:** 10.64898/2026.05.22.26353861

**Authors:** Josh King-Robson, Molly R E Cartlidge, Eyal Soreq, Heidi Murray-Smith, Matthew Harrison, Sophie Horrocks, Lina Aimola, Marie Poole, Ríona Mc Ardle, Louise Robinson, David J Sharp, Jonathan M Schott

**Affiliations:** Dementia Research Centre, UCL Queen Square Institute of Neurology, University College London, London, UK; UK Dementia Research Institute, University College London, London, UK; UCL Consultants, London, UK; UK Dementia Research Institute Centre for Care Research & Technology, Imperial College London and University of Surrey, London, UK; Department of Brain Sciences, Imperial College London, London, UK; Sorpol Consultancy, Ashdod, Israel; Department of Surgery & Cancer, Faculty of Medicine, Imperial College London, UK; Population Health Sciences Institute, Newcastle University, Newcastle, UK; Translational and Clinical Research Institute, Newcastle University, Newcastle, UK; NIHR Newcastle Biomedical Research Centre, Newcastle University, Newcastle, UK

**Keywords:** Sleep, Dementia, biomarkers, Digital health, Protocol, Feasibility study

## Abstract

**Background:** Improvements in health technology offer opportunities for remote disease screening, diagnosis and monitoring. The Withings Sleep Analyzer (WSA), an under mattress ballistocardiograph sensor able to detect body movement, breathing, and cardiac ejection is a promising technology for the non-invasive detection and monitoring of neurodegenerative diseases. InSleep46 aims to evaluate whether the WSA is able to detect preclinical Alzheimer’s disease in members of the 1946 British Birth cohort, now in their late 70s.

**Objectives:** To assess feasibility of deployment of a remote sleep, circadian and physiological monitoring device in a population of older adults.

**Participants:** 356 participants from the Insight 46 neuroimaging sub-study (1946 British Birth Cohort), all born in one week in March 1946.

**Methods:** We describe remote recruitment, device installation, and troubleshooting protocols. Feasibility analysis examined participant characteristics associated with recruitment and successful device setup using logistic regression. Troubleshooting events for device installation and maintenance were recorded over a mean 14-month follow-up period.

**Results:** During the feasibility analysis period, 263 (74%) participants, mean (SD) age 77 years (0.47) agreed to take part, of whom 245 (93%) successfully set up the WSA. Recruitment and successful set up of the WSA were not dependent on cognitive ability, socioeconomic position, or educational attainment. 162 (62%) of recruited individuals required ≥1 troubleshooting call (mean 2.3 per participant, range 0-16). 603 calls were required in total.

**Conclusion:** Deployment of a remote sleep and physiological monitoring device in an older adult population is feasible. Most participants required individualised assistance to set up the device. For the technology to be widely implemented, the set up must be accessible, with dedicated support available.

**STRENGTHS AND LIMITATIONS OF THIS STUDY:**

- The Withings Sleep Analyzer, a ballistocardiographic under-mattress sensor, enables passive collection of sleep, circadian, and physiological data in participants’ own homes, maximising ecological validity with minimal demand on research participants.
- Study participants are from the world’s longest continuously running birth cohort, with extensive longitudinal data available, collected over 27-waves of data collection from birth to age 80, including cognitive, imaging and fluid biomarkers for neurodegenerative disease.
- This is a nationally representative cohort of participants who are all the same age. However, reflecting the population of post-war Britain at its inception, the cohort is not ethnically diverse and thus findings may not be generalisable to all ethnic groups. Furthermore, as those invited have/expect to attend a two-day clinic visit, the cohort is likely to consist of more willing and able participants than the general population.
- This study prospectively collected data on the feasibility of deployment and set up of a remote sleep monitoring device, enabling understanding of potential barriers to widescale deployment of remote digital biomarkers.
- We provide a practical framework for the successful deployment and maintenance of a remote digital monitoring device.

## INTRODUCTION

Technology for remote diagnosis and disease monitoring is increasingly prevalent in clinical practice, including the management of chronic conditions such as diabetes or heart failure, and for gold-standard diagnosis of hypertension.^1-3^ With an aging population, remote monitoring devices offer opportunities to screen, diagnose, and monitor health conditions impacting older adults, such as Alzheimer’s disease.^4 5^

### Use of health technology in monitoring cognitive performance and diagnosing dementia

A wide range of diagnostic tools are under evaluation for their ability to identify and monitor those with Alzheimer’s disease and other dementias. The emergence of sensitive blood biomarkers for Alzheimer’s disease pathology offers opportunities for early pathological diagnosis; however, these markers can be unreliable where common comorbidities are present.^6^ Due to the long prodromal period of pathological protein accumulation in Alzheimer’s disease, blood biomarkers are limited in their ability to predict proximity to symptomatic onset, which can complicate the interpretation of results.^7^ Remote digital cognitive testing can detect cognitive decline associated with imaging markers of neurodegeneration;^8^ however, as Alzheimer’s disease pathology is present years prior to incipient cognitive change,^9^ there is a need for the development of biomarkers that can detect and monitor symptoms of pre-clinical disease. This need has been emphasized by the National Health Service (NHS) 10-year plan, and a call by the World Health Organization for the use of innovative health technologies in the early diagnosis of dementia.^10 11^

### Digital biomarkers for monitoring sleep and remotely assessing dementia risk

Sleep and circadian disturbances are common features of neurodegenerative disease.^12 13^ Cognitively normal individuals with sleep disturbances have a higher risk of developing Alzheimer’s disease, vascular dementia and all-cause dementia over the subsequent two decades,^14^ raising the prospect that a digital sleep biomarker could help to identify those at increased dementia risk. However, the exact sleep and circadian features that predict or detect incipient neurodegeneration remain unclear. Current ‘gold standard’ polysomnography is expensive and demands considerable time and effort from patients, making it impractical for long-term monitoring, while testing in a laboratory environment limits ecological validity.^7^ There is a need to develop new, cost-effective methods to measure the sleep of patients in the community. At-home monitoring, including the use of ballistocardiographic under-mattress devices, has been successfully used to assess the health of individuals in the community.^5 15^

The Withings Sleep Analyzer (WSA); a ballistocardiographic under-mattress device, is able to detect body movements, chest displacement (breathing), and vibrations due to cardiac ejection from which metrics of bed occupancy, sleep, respiration, heart rate and variability can be derived.^5^ A microphone enables snoring detection. The WSA has been validated for the detection of obstructive apnoea^16^. The Dementia Research Institute Sleep Index for AD (DRI-SI-AD), a digital biomarker quantifying sleep disturbances from data collected using the WSA, can identify dementia related sleep disturbances and accurately differentiates people living with dementia from age matched controls.^5^ It is unclear whether a digital sleep biomarker can identify pre-clinical AD pathology.

InSleep46 aims to develop a digital sleep, circadian and physiological biomarker for the detection of AD pathology in pre-clinical populations. Participants are recruited from the Insight 46 neuroscience substudy of the world’s longest continuously running birth cohort and have a wide range of cognitive, imaging and fluid biomarkers for neurodegenerative disease available. It is necessary to understand the applicability of remote sleep and physiological monitoring in older patients’ homes to assess dementia risk in preclinical phases, and it remains unclear whether the remote deployment of these devices is feasible in research or clinical contexts. We share a practical framework for the successful deployment of a remote sleep, circadian and physiological monitoring device in a cohort of older adult individuals in a community setting, and we examine the feasibility and resource requirements.

## METHODS AND ANALYSIS

### Study organization

#### NSHD and Insight 46

The MRC National Survey of Health and Development (NSHD) is a longitudinal study following the

1946 British Birth Cohort. Originally comprising 5,362 individuals born in Great Britain in the first week of March in 1946, participants have now completed 27 waves of data collection over 80 years of follow-up.^17^ In addition to the cohort-wide data collection, multiple sub-studies have been conducted, including Insight 46 (a neuroimaging sub-study of the NSHD). The protocol for Insight 46 has been described previously.^18 19^ In summary, 502 NSHD study members were recruited at age ∼70 to undergo serial neuroscience assessment (including neuroimaging, fluid biomarker, cognitive assessment) in London. These participants were invited to return for a visit at age ∼73, and at a third time point at age ∼77 for repeat (longitudinal) assessments. Data collection at age ∼77 was supplemented by an additional 300 NSHD members who had not previously attended any Insight 46 research visit.

#### InSleep46

InSleep46 was designed with the aim to test a digital biomarker for nighttime behaviour and sleep in a longitudinal cohort. This protocol builds upon experience gained from ongoing remote assessments using the WSA and in the Insight 46 cohort.^8 19^ Data collection is due to complete in August 2028.

### Patient and public involvement

NSHD participants are regularly involved in the design of the study through a range of engagement activities and focus groups with every effort made to ensure wide and equitable participation independent of personal circumstances and means.

This project was complimented with a parallel work package to explore the public and professional acceptability of using sleep sensors as part of a pre-diagnostic service, through four public consultation workshops (n=31) and interviews with general practitioners (n=8). A qualitative research approach was used to analyse this work.^20^ Over the course of the project, two lay advisors with personal experience of a dementia diagnosis reviewed the funding application, helped shape consultation work, attended cross-project meetings, and reviewed and commented on project outputs.

### Recruitment

The overarching aim of this study is to investigate the feasibility of continuously acquired measures of sleep and circadian function in the home as a biomarker for detection of preclinical dementia and proximity to cognitive decline. Sample sizes were calculated utilising 180,000 nights of data from the WSA collected from people of similar age to Insight 46 participants who had >180 nights of data available alongside Pittsburgh Sleep Quality Index (PSQI). Our initial recruitment target of 250 participants was powered to identify small differences in sleep quality as measured using the PSQI (>0.2-unit group differences) between amyloid +ve/-ve groups with a power of 0.95 and alpha of 0.05. We have invited a total of 356 participants (**Figure 2A**).

Recruitment for the InSleep46 project is ongoing. Insight 46 participants were invited to take part in InSleep46 if they completed or were expected to complete a third Insight 46 research visit. As of December 2024, 297 participants who had completed longitudinal Insight 46 assessments had been invited to take part in InSleep46, supplemented by 59 participants who were invited having taken part in only a single Insight 46 visit (**Figure 2A**).

Participants were sent an invitation in the post containing a participant information sheet and an InSleep46 consent form. They were called by a member of the study team to confirm interest, eligibility, that minimum technological requirements were met, and consent (**Supplementary Figure 1**). If the participant was eligible to take part, they were asked a series of questions regarding their sleeping habits (**Supplementary Table 1**).

Participants were given the option to complete the consent form in one of two ways—either by completing a paper copy and returning it in the post to the study team, or by completing an online version collected and managed using REDCap electronic data capture tools hosted at University College London (UCL). REDCap (Research Electronic Data Capture) is a secure, web-based software platform designed to support data capture for research studies, providing 1) an intuitive interface for validated data capture; 2) audit trails for tracking data manipulation and export procedures; 3) automated export procedures for seamless data downloads to common statistical packages; and 4) procedures for data integration and interoperability with external sources.^21 22^ In both cases, the member of the study team undertaking consent would read through the consent form with the participant over the phone and answer any questions the participant had about the study or data collection prior to the participant providing their informed consent.

### Deployment

Upon receipt of the completed consent form, each study member was sent a WSA, alongside written and video instructions (**Figure 1**, https://mindermeetingplace.com/insleep46/). If the participant did not have Wi-Fi or access to a smart phone or tablet, they were also sent a cellular ‘data hub’ modem.

**Figure 1:**
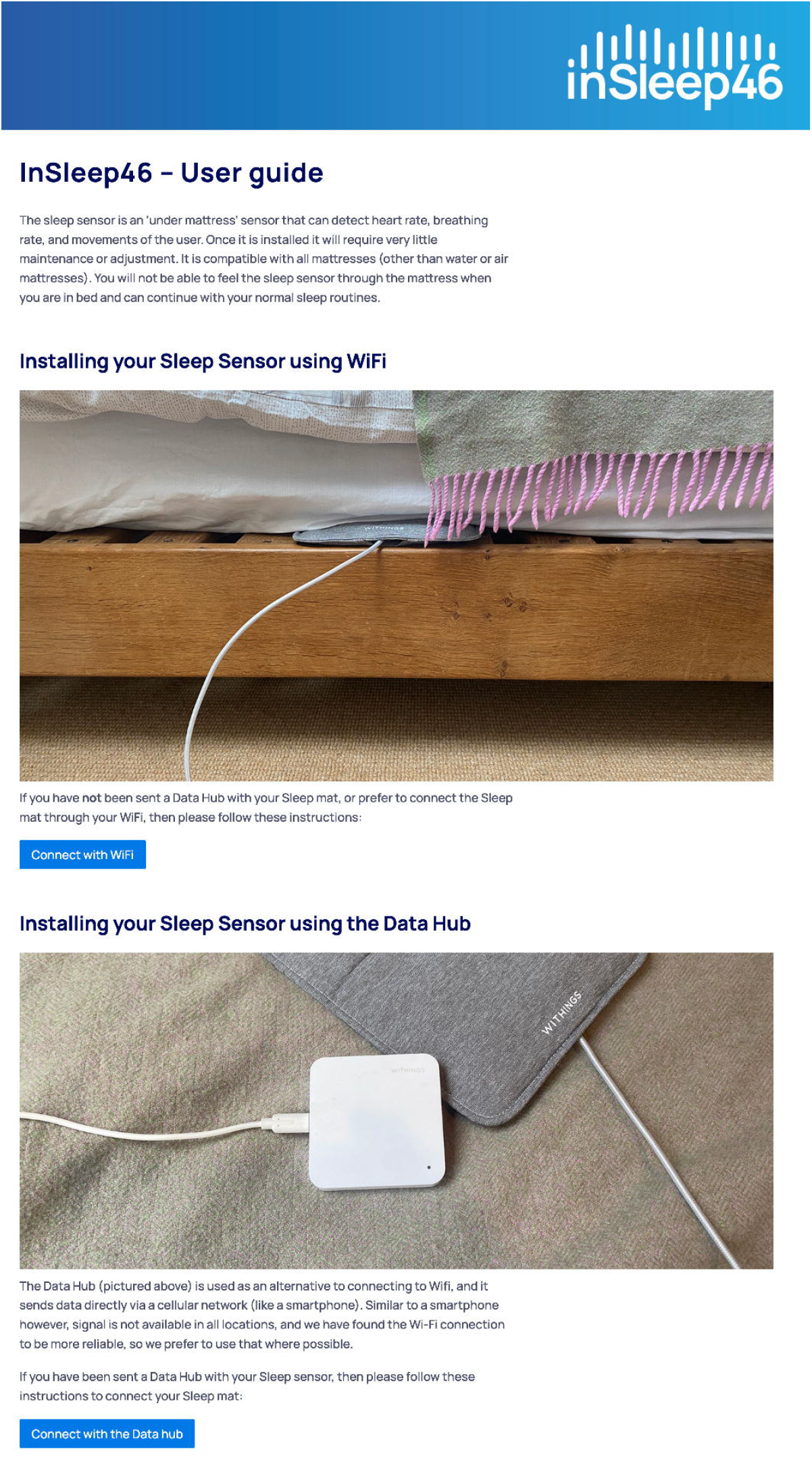
Devices and remote support. The InSleep46 welcome page (https://mindermeetingplace.com/insleep46/) provides technical support, including step by step videos for device installation and common technical issues. The Withings Sleep Analyzer (WSA, top) is shown positioned under a mattress, alongside a data hub (bottom).

#### Data flow and Minder

Data from the WSA were transferred either via Wi-Fi or cellular signal to Withings and stored on secure servers in Europe (GDPR and ISO27001 certified). Data were then transferred to secure servers at the UK Dementia Research Institute (UK DRI) Care, Research and Technology Centre (CR&T) based at Imperial College London. Once the InSleep46 participant connected the WSA to their home Wi-Fi or data hub, data were viewable contemporaneously by approved members of the study team on the Minder platform—developed at the CR&T—which enabled the team to confirm data flow.

### Troubleshooting

Participants were asked to connect the WSA within two weeks of receiving it. If data were not received within a month of deployment, the participant was contacted by the study team to identify the cause of delay and provide support (**Supplementary Table 2**). A troubleshooting entry was logged for each troubleshooting instance. Study participants were also able to email or call a dedicated helpline for assistance from the UCL research team.

### Feasibility analysis

Prospective collection of data at all stages of recruitment and deployment of the WSA enabled evaluation of feasibility and barriers to successful deployment of a remote digital biomarker in older adult populations. The feasibility study analysis plan was developed based on extensive prior experience of data collection with the NSHD cohort and informed by prior analysis of recruitment rates to the cohort’s sub-studies including Insight 46,^23^ and feasibility studies for other remote assessments.^24^

#### Troubleshooting and outcome data

Feasibility was assessed by calculating the proportion of participants who were recruited and successfully set up the WSA, delays in reaching these milestones, the total number of nights of data collected, and the completeness of this data (percentage of nights of data collected out of maximum possible since WSA deployment).

All logged troubleshooting instances (>1 hour apart) were categorised into eleven categories based on the primary subject of the call: participant unavailable, participant holiday/sleeping pattern change, participant medical issue, participant passed away, postage or administrative error, reporting successful set up, social issue, participant withdrew from study (either before or after successful connection), technical issue–hub, technical issue–Wi-Fi, and technical issue–Sleep Analyzer/general. Troubleshooting instances could be initiated by study participants or the study team.

#### Predictors of participation

We investigated the relationship between sociodemographic and health-related characteristics and the likelihood of an invited individual being eligible and consenting to take part in InSleep46, and the subsequent likelihood of them successfully connecting their WSA.

Educational attainment was determined as the highest qualification achieved by age 26, categorised into three groups: no qualification, up to O-level or equivalent (secondary school), and A-level (higher secondary/college) equivalent or higher. Childhood socioeconomic position (SEP) was defined using the paternal SEP when the participant was aged 11 (or 15 or 4 where unavailable), according to the Office of Population Censuses and Surveys classification, and categorised into manual or non-manual. Adulthood SEP was derived from the individual’s own occupation at 53, using the same methodology. Childhood cognition was measured at age 8 (or age 11/15 where unavailable) using four tests of verbal/non-verbal ability, converted into a z-score based on the full NSHD cohort.^25^ Adult cognition was determined using the Preclinical Alzheimer Cognitive Composite (PACC) at age ∼77, converted into a z-score based upon the mean and standard deviation of scores in the Insight 46 cohort at age ∼70. The PACC combines the Mini Mental State Examination, Logical Memory IIa from the Wechsler Memory Scale-Revised, Digit-Symbol Substitution test from the Wechsler Adult Intelligence Scale-Revised, and 12-item Face-Name test into a score sensitive to preclinical cognitive decline.^25^ Lifetime smoking was self-reported at age 76 and recoded into never smoked, ex-smoker and current smoker. Alcohol use was self-reported at age 76 and recoded as never, up to once per week, 2-3 times per week, and 4 or more times per week. Self-rated health at age 76 was recoded into three categories: fair or poor, good, and very good or excellent. Functional independence was self-reported at age 76 according to need for assistance with feeding, grooming, using the stairs, using the toilet, or moving on level surfaces. APOE ⍰4 carrier status was determined from blood sampling at age 53 and grouped into non-carrier or carrier (heterozygous or homozygous).

#### Statistical analysis

Logistic regression estimated associations between each potential predictor of participation, both successful recruitment and data connection. Models were initially unadjusted, and subsequently adjusted for sex, educational attainment, childhood, and adult SEP. Coefficients are exponentiated to present odds ratios. Values of p<0.05 are considered statistically significant for all analyses.

## RESULTS

### Recruitment and participant characteristics

As of December 2024, 356 participants were invited to take part in InSleep46 (**Table 1, Figure 2A**). Of these, 263 participants (74%) were recruited and sent a WSA, and 245 participants (93%) successfully connected their WSA, over a mean (SD) 14 (4) month follow-up period from initial invitation.

**Table 1.**
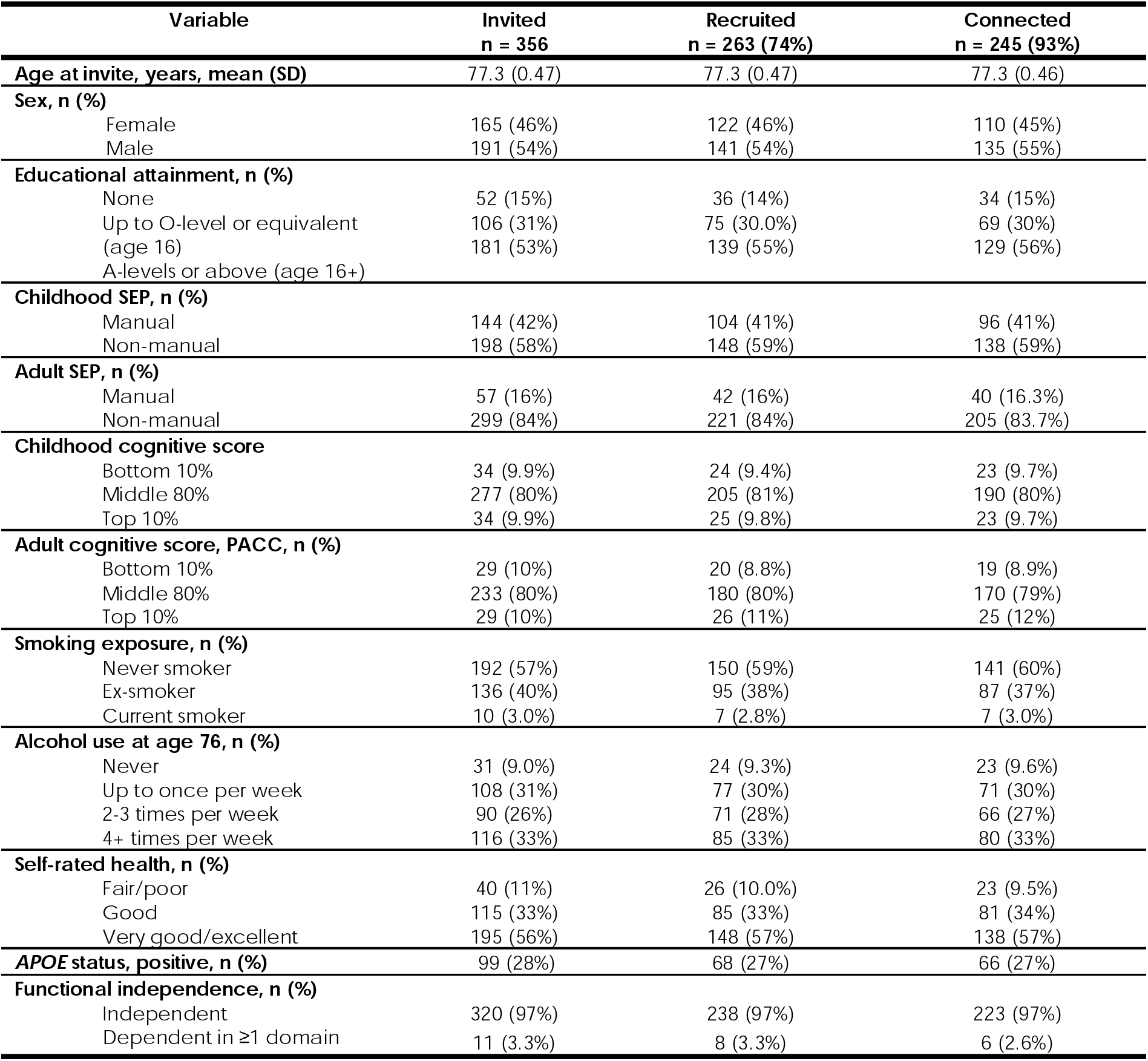
Socioeconomic and health characteristics for those invited to, recruited to, and successfully connected their WSA in InSleep46. SD; standard deviation, SEP; socioeconomic position, PACC; Preclinical Alzheimer Cognitive Composite. Data were unavailable for Childhood SEP (n=14), childhood cognition (n=11), PACC (n=65), smoking exposure (n=18), alcohol use (n=11), self-rated health (n=6), APOE status (n=7) and functional independence (n=25).

| Variable | Invited<br>n = 356 | Recruited<br>n = 263 (74%) | Connected<br>n = 245 (93%) |
| --- | --- | --- | --- |
| **Age at invite, years, mean (SD)** | 77.3 (0.47) | 77.3 (0.47) | 77.3 (0.46) |
| **Sex, n (%)** |  |  |  |
| Female | 165 (46%) | 122 (46%) | 110 (45%) |
| Male | 191 (54%) | 141 (54%) | 135 (55%) |
| **Educational attainment, n (%)** |  |  |  |
| None | 52 (15%) | 36 (14%) | 34 (15%) |
| Up to O-level or equivalent<br>(age 16) | 106 (31%) | 75 (30.0%) | 69 (30%) |
| A-levels or above (age 16+) | 181 (53%) | 139 (55%) | 129 (56%) |
| **Childhood SEP, n (%)** |  |  |  |
| Manual | 144 (42%) | 104 (41%) | 96 (41%) |
| Non-manual | 198 (58%) | 148 (59%) | 138 (59%) |
| **Adult SEP, n (%)** |  |  |  |
| Manual | 57 (16%) | 42 (16%) | 40 (16.3%) |
| Non-manual | 299 (84%) | 221 (84%) | 205 (83.7%) |
| **Childhood cognitive score** |  |  |  |
| Bottom 10% | 34 (9.9%) | 24 (9.4%) | 23 (9.7%) |
| Middle 80% | 277 (80%) | 205 (81%) | 190 (80%) |
| Top 10% | 34 (9.9%) | 25 (9.8%) | 23 (9.7%) |
| **Adult cognitive score, PACC, n (%)** |  |  |  |
| Bottom 10% | 29 (10%) | 20 (8.8%) | 19 (8.9%) |
| Middle 80% | 233 (80%) | 180 (80%) | 170 (79%) |
| Top 10% | 29 (10%) | 26 (11%) | 25 (12%) |
| **Smoking exposure, n (%)** |  |  |  |
| Never smoker | 192 (57%) | 150 (59%) | 141 (60%) |
| Ex-smoker | 136 (40%) | 95 (38%) | 87 (37%) |
| Current smoker | 10 (3.0%) | 7 (2.8%) | 7 (3.0%) |
| **Alcohol use at age 76, n (%)** |  |  |  |
| Never | 31 (9.0%) | 24 (9.3%) | 23 (9.6%) |
| Up to once per week | 108 (31%) | 77 (30%) | 71 (30%) |
| 2-3 times per week | 90 (26%) | 71 (28%) | 66 (27%) |
| 4+ times per week | 116 (33%) | 85 (33%) | 80 (33%) |
| **Self-rated health, n (%)** |  |  |  |
| Fair/poor | 40 (11%) | 26 (10.0%) | 23 (9.5%) |
| Good | 115 (33%) | 85 (33%) | 81 (34%) |
| Very good/excellent | 195 (56%) | 148 (57%) | 138 (57%) |
| **APOE status, positive, n (%)** | 99 (28%) | 68 (27%) | 66 (27%) |
| **Functional independence, n (%)** |  |  |  |
| Independent | 320 (97%) | 238 (97%) | 223 (97%) |
| Dependent in ≥1 domain | 11 (3.3%) | 8 (3.3%) | 6 (2.6%) |

**Figure 2:**
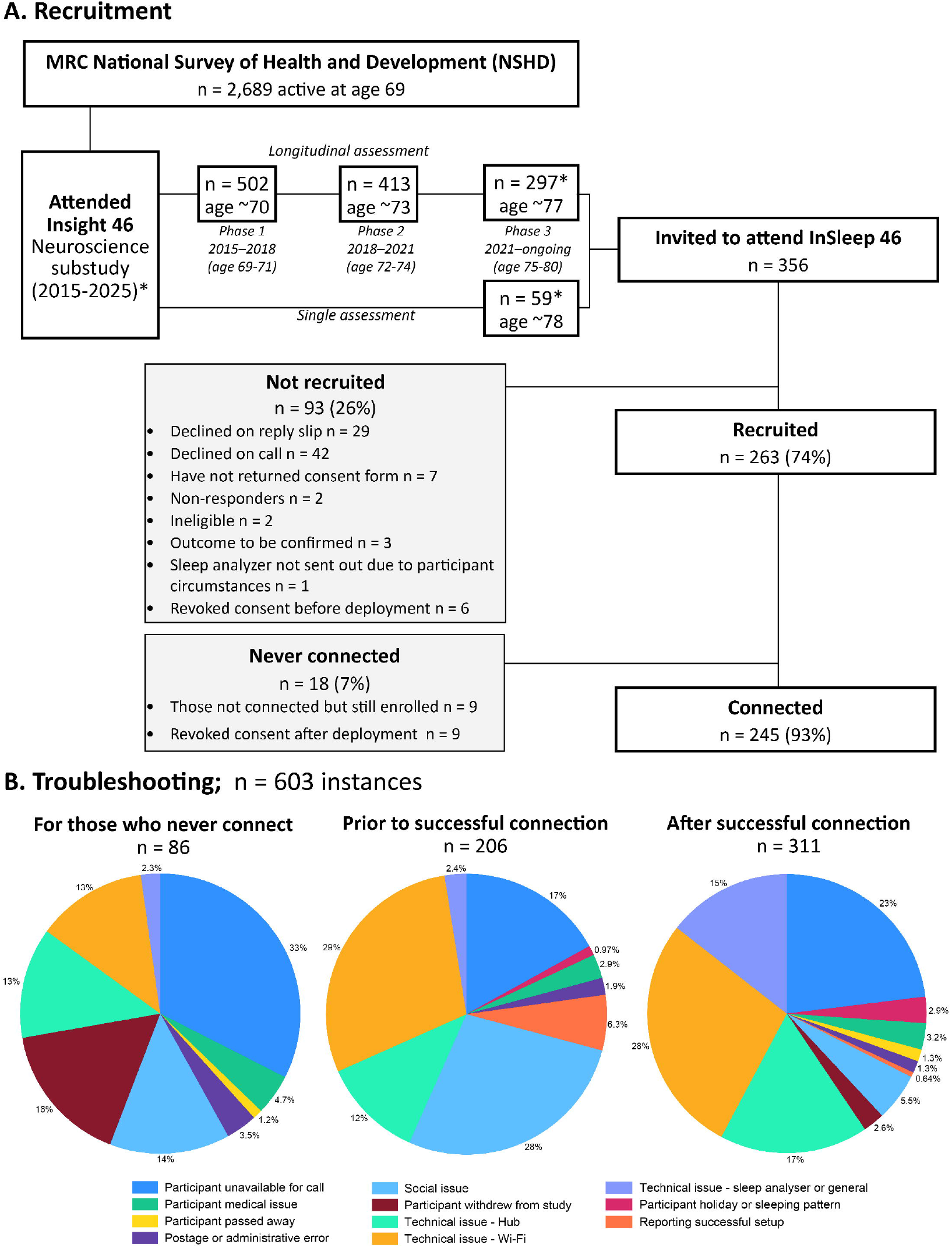
Recruitment and deployment to InSleep46. **A**: Flowchart showing stages from invitation to successful Withings Sleep Analyzer (WSA) connection. **B**: The main subject for each troubleshooting instances, separated into three groups; those for who participants who never connected, those that occurred before connection, and those which occurred after connection. Insight 46 recruitment is ongoing.*

Reasons for non-recruitment are provided in **Figure 2B**. For those who were sent a WSA, the median time to set up their WSA from arrival (assuming two-day postage), was 8 days (range from 0-313). Of those participants who never connected, n=9 revoked consent and n=9 still planned to set up their WSA. Those who revoked consent have done so for the following reasons: participant passed away (n=1), health concerns (n=2), spouse’s concerns over technology (n=1), lost interest after technical issues (n=2), and reasons not specified (n=3).

### Data completeness

64,834 nights of data were collected by 22^nd^ February 2025. The number of participants connected increased steadily across a maximum 18-month follow-up period (**Figure 3**). The mean data completeness (percentage of nights data collected out of maximum possible since WSA deployment) was 83.3% (SD 17%, **Figure 4**).

**Figure 3:**
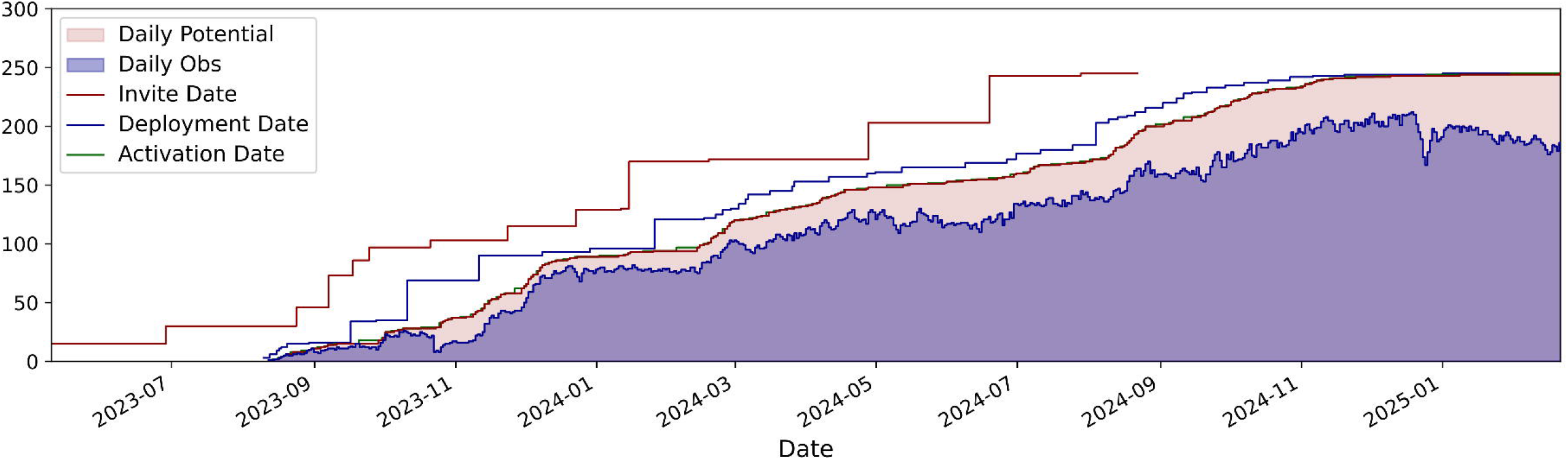
Data accrual. Step curves show the cumulative number of participants across key milestones: invite (red line, up to August 2024), deployment (blue line), and activation (green line). The shaded areas depict the daily observed data (blue); and potential (maximum possible) data collected.

**Figure 4:**
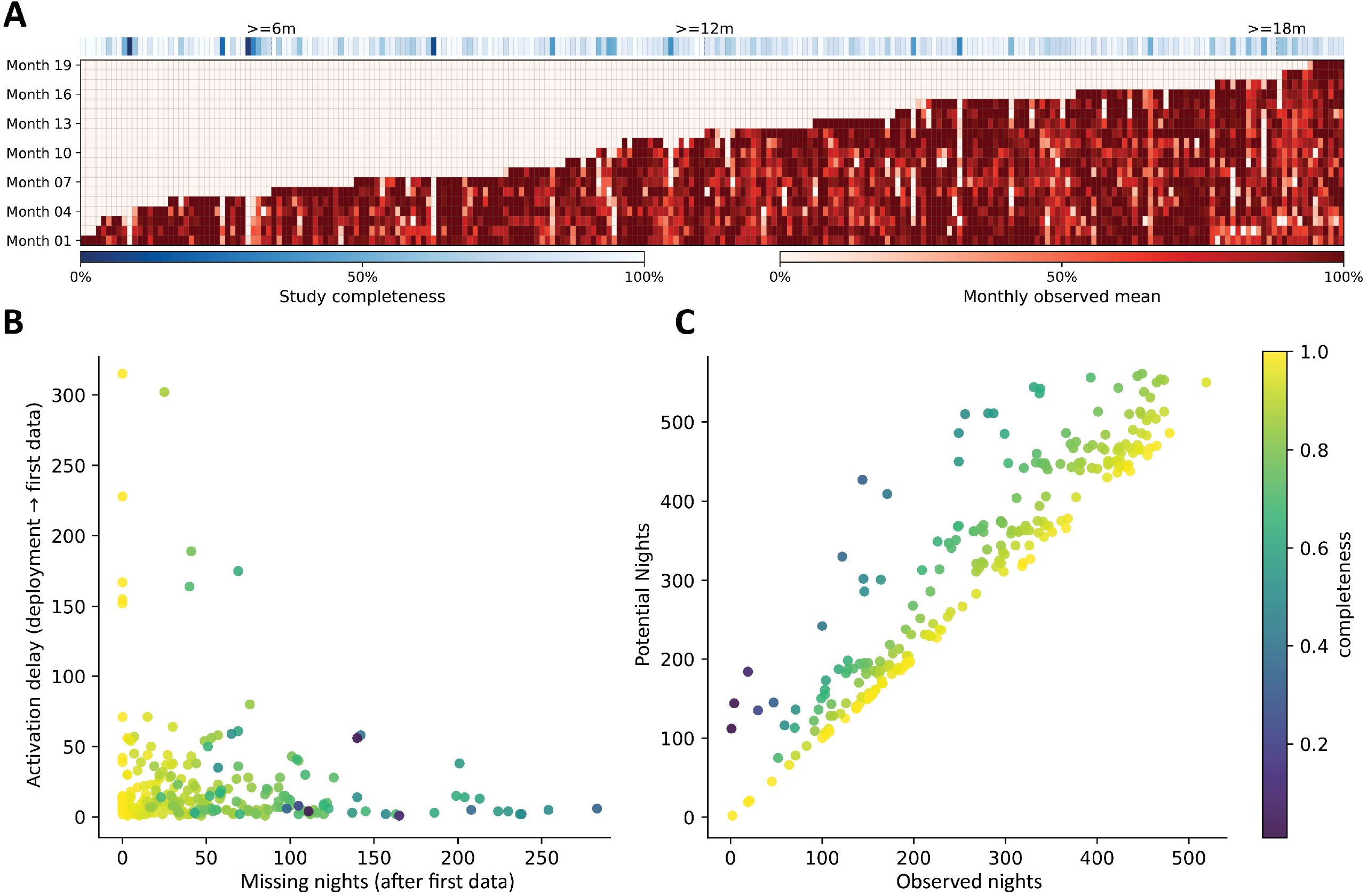
Data completeness. **A:** Heatmap showing data collected per participant. Each column represents one participant, ordered by date of device deployment (left=most recent). Rows represent months since each participant’s commencement in the study (first month at the bottom). Shades of red encode the mean data completeness during each month (dark red = 100%, white = 0%). The top colour bar represents each participant’s overall data completeness since enrolment in the study (white = 100%, dark blue = 0%). The scatterplots summarize **B**: time from device deployment to successful connection vs number of missing nights after successful connection, and C: observed vs potential nights of data collected. Points are coloured by each participant’s overall data completeness.

### Troubleshooting

There were 603 recorded troubleshooting instances within the 14-month follow-up period (**Figure 2B**); the mean number of troubleshooting calls was 2.3 per participant (range 0-16) among 263 participants recruited and sent a WSA. 101 participants (38%) did not require any troubleshooting assistance at any time. Of those 162 participants (62%) who required troubleshooting during the 14-month follow-up period, 78 (30%) required assistance prior to connection, 119 (45%) required assistance after connection, and 86 (33%) required assistance both before and after connection. The 18 participants (6.8%) who never successfully connected all required ≥1 troubleshooting call.

292 (48%) of troubleshooting instances were logged prior to obtaining a connection; 86 (14%) were for participants who had a WSA distributed but never successfully connected, 206 (34%) were for participants in whom troubleshooting preceded a successful connection, and 311 (52%) were logged after obtaining a successful connection (**Figure 2B**). **Supplementary Table 2** outlines typical issues from each troubleshooting category and steps taken/advised for their resolution.

### Predictors of participation

**Table 1** shows the distribution of each potential predictor of recruitment in relation to invitation, recruitment, and successful connection of the WSA. The mean (SD) age at invite was 77.3 (0.47) years, 54% of those invited were male, and 3.3% of those invited were dependent in ≥1 functional domain.

Predictors of recruitment to InSleep46 and subsequent successful WSA set-up are presented in **Table 2**. None of the characteristics investigated was found to significantly influence the likelihood of recruitment or successful connection. However, there was a non-significant trend toward increased rate of recruitment in those with the top 10% of adult cognitive scores compared to those with the bottom 10% (odds ratio (OR) 3.90, 95% CI 0.93 to 16.3, p=0.06), and a lower rate of successful connection in female participants (OR 0.41, 95% CI 0.15 to 1.12, p=0.08) and in participants with functional dependence in ≥1 domain (OR 0.20, 95% CI 0.04 to 1.09, p=0.06).

**Table 2:** Analyses of socioeconomic and health characteristic predictors for InSleep46 recruitment and WSA connection. Results of logistic regression analyses examining factors predictive of InSleep46 recruitment and Withings Sleep Analyzer (WSA) connection, adjusted for sex, education attainment, child and adult socioeconomic position (SEP). Analyses of sex, education attainment, child and adult SEP were adjusted for the three remaining variables. OR, odds ratio; CI, confidence interval, PACC; Preclinical Alzheimer Cognitive Composite.

| Variable | Recruited (max n = 356) |  |  | Connected (max n = 263) |  |  |
| --- | --- | --- | --- | --- | --- | --- |
|  | OR | p | 95% CI | OR | p | 95% CI |
| **Sex** |  |  |  |  |  |  |
| Male | Reference |  |  | Reference |  |  |
| Female | 1.01 | 1.0 | 0.63, 1.62 | 0.41 | 0.08 | 0.15, 1.12 |
| **Childhood SEP** |  |  |  |  |  |  |
| Manual | Reference |  |  | Reference |  |  |
| Non-manual | 1.14 | 0.6 | 0.70, 1.85 | 1.15 | 0.8 | 0.44, 3.02 |
| **Educational attainment** |  |  |  |  |  |  |
| None | Reference |  |  | Reference |  |  |
| Up to O-level or equivalent (age 16) | 1.08 | 0.8 | 0.52, 2.21 | 0.67 | 0.6 | 0.13, 3.53 |
| A-level and above (age 16+) | 1.47 | 0.3 | 0.74, 2.91 | 0.76 | 0.7 | 0.16, 3.63 |
| **Adult SEP** |  |  |  |  |  |  |
| Manual | Reference |  |  | Reference |  |  |
| Non-manual | 1.01 | 1.0 | 0.53, 1.93 | 0.64 | 0.6 | 0.14, 2.90 |
| **Childhood cognitive score** |  |  |  |  |  |  |
| Bottom 10% | Reference |  |  | Reference |  |  |
| Middle 80% | 1.19 | 0.7 | 0.54, 2.60 | 0.55 | 0.6 | 0.07, 4.36 |
| Top 10% | 1.16 | 0.8 | 0.40, 3.34 | 0.50 | 0.6 | 0.04, 5.91 |
| **Adult cognitive score, PACC** |  |  |  |  |  |  |
| Bottom 10% | Reference |  |  | Reference |  |  |
| Middle 80% | 1.53 | 0.3 | 0.66, 3.56 | 0.89 | 0.9 | 0.11, 7.38 |
| Top 10% | 3.90 | 0.06 | 0.93, 16.31 | 1.32 | 0.8 | 0.08, 22.4 |
| **Smoking exposure** |  |  |  |  |  |  |
| Never smoked | Reference |  |  | Reference |  |  |
| Current or ex-smoker | 0.65 | 0.09 | 0.40, 1.06 | 0.75 | 0.6 | 0.28, 2.01 |
| **Alcohol use at age 76** |  |  |  |  |  |  |
| Never | Reference |  |  | Reference |  |  |
| Up to once per week | 0.72 | 0.5 | 0.28, 1.85 | 0.51 | 0.6 | 0.06, 4.50 |
| 2-3 times per week | 1.09 | 0.9 | 0.41, 2.91 | 0.57 | 0.6 | 0.06, 5.17 |
| 4+ times per week | 0.80 | 0.6 | 0.31, 2.04 | 0.70 | 0.8 | 0.08, 6.23 |
| **Self-rated health at age 76-77** |  |  |  |  |  |  |
| Fair/poor | Reference |  |  | Reference |  |  |
| Good | 1.53 | 0.3 | 0.71, 3.30 | 2.64 | 0.2 | 0.55, 12.66 |
| Very good/excellent | 1.70 | 0.2 | 0.82, 3.51 | 1.80 | 0.4 | 0.46, 7.04 |
| **APOE status** |  |  |  |  |  |  |
| Negative | Reference |  |  | Reference |  |  |
| Positive (heterozygous or homozygous) | 0.71 | 0.2 | 0.42, 1.18 | 2.64 | 0.2 | 0.58, 11.93 |
| **Functional independence** |  |  |  |  |  |  |
| Independent | Reference |  |  | Reference |  |  |
| Dependent in ≥1 domain | 0.92 | 0.9 | 0.24, 3.55 | 0.20 | 0.06 | 0.04, 1.09 |

The results of the same models adjusted for sex, educational attainment, child and adult SEP are presented in **Supplementary Table 3**.

## DISCUSSION

We demonstrate that the deployment of a remote sleep and circadian monitoring device in a population-based cohort in their late 70s is feasible, and we outline a framework for its implementation. Remote deployment of these devices offers potential opportunities for diagnosis, disease monitoring, and population-based screening. We successfully collected almost 65,000 nights of data during our feasibility study, with data collected from 93% of recruited participants; however, this required individualised troubleshooting support for over half of the study participants.

A total of 603 troubleshooting calls, averaging 2.3 calls (range 0-16) per participant were required over a mean (SD) 14 (4) month follow-up period, with 59% of successfully connected participants requiring individualised support. Many troubleshooting calls were related to technological barriers. While some of these were specific to the WSA, there are themes which would be applicable to other remote technologies, such as the need to connect a device to Wi-Fi or a cellular data connection. Individuals’ ability to use technology has been shown to be a barrier to the use of remote testing in other studies. Remote cognitive assessments within the NSHD cohort also reported that the majority of calls to their helpline focused on technical queries^24^ related to accessing the online consent and technical issues with the platform itself.

Social issues, herein including the need for support by family/friends, were a further barrier to successful set up of the device. This has been found in other studies of health technology, with 82% of older adults accessing video home-based primary care during the COVID-19 pandemic requiring support to use the technology.^26^ Limited technological literacy and the prominence of medical comorbidities, which both delayed and prohibited the setup of the WSA in our study, are more prevalent in older populations.^27 28^ There is a need to ensure that the technological requirements of systems planned for use by older adults are as user-friendly as possible to prevent digital exclusion from widening existing health inequalities.^29^

To successfully set up their WSA, InSleep46 participants required either a stable Wi-Fi connection with access to a smart phone or tablet, or sufficient cellular signal within their bedroom to use the data hub. Individuals were ineligible for recruitment if they did not fulfil either of these requirements. Stronger cellular signals across the UK, especially in rural areas, would be needed to roll out devices such as the WSA as a widely used diagnostic tool so that a lack of Wi-Fi connection is not a barrier which further exacerbates digital exclusion.

This work found that participants were able to successfully set up the WSA, regardless of cognitive ability, socioeconomic position, and educational attainment. Although we note a non-significant trend towards lower rates of successful deployment in females and those with reduced functional independence, rates of successful connection remained high in all groups. There are notable limitations to these findings; specifically, NSHD participants are all the same age and the cohort lacks racial or ethnic diversity, both factors known to influence the adoption of healthcare technology.^30^ Further study is required to confirm if these devices can be successfully deployed in populations outside of the UK, given the marked differences in technology use across global populations.^29^ Furthermore, InSleep46 participants had already agreed to attend a two-day in person visit including a number of cognitively and physically demanding assessments, with the majority travelling to London from across the UK for assessment,^19^ and are therefore likely healthier and more physically able than the general population.

### Plans for future analysis

Having established feasibility, future analyses will focus on developing and validating a digital sleep biomarker for preclinical neurodegeneration. We will evaluate the ability of the DRI-SI-AD to detect preclinical AD pathology (β-amyloid and tau) as well as developing new modelling approaches for the preclinical detection of brain pathologies, prioritising interpretability and clinically deployable approaches to analyse the large volume of data produced, aiming to balance parsimony, predictive performance, and transparency. Development will follow established best practice, including appropriate data partitioning, internal validation, and evaluation of discrimination, calibration, and potential clinical utility to support generalisability and real-world application. Our ultimate aim is to provide a non-invasive reliable method for determining which elderly individuals are at imminent risk of developing cognitive symptoms due to Alzheimer’s disease, to inform prevention trials and ultimately provide a scalable means of population-based screening.

## Conclusion

We demonstrate that deployment of a remote sleep and circadian monitoring device in a population-based cohort in their late 70s is feasible; however, more than half of individuals required individualised support to connect the WSA. This work highlights the importance of ensuring remote devices used to detect and monitor health conditions in older adult populations are accessible and that sufficient support is available. We share our methodology for successful deployment of this and similar devices.

## Ethics and Dissemination

This study was approved by the Health Research Authority Research Ethics Committee (HRA REC) London (REC reference 14/LO/1173, PI Schott). All participants provided written informed consent to participate before they were sent a WSA, in accordance with the Declaration of Helsinki, and for their data to be stored in accordance with the Data Protection Act (2018) and General Data Protection Regulations (GDPR).

Results will be submitted for publication in peer-reviewed journals and presented at conferences.

## Data availability

The NSHD data-sharing policy is available on the NSHD Data Sharing website (https://nshd.mrc.ac.uk/data-sharing). Data generated as part of Insight 46, including inSleep46, will be made available one year after data collection is complete, allowing sufficient time for quality control. Data will be made available to bona fide researchers through UCL’s Unit for Lifelong Health and Ageing’s standard security process, and in accordance to established NSHD data sharing guidelines (http://www.nshd.mrc.ac.uk/data). doi: 10.5522/NSHD/Q101, doi: 10.5522/NSHD/Q102, and doi: 10.5522/NSHD/Q103.

## Acknowledgements

We are very grateful to the study participants for their contributions to InSleep46 and for their commitments to research over the last eight decades, and the NSHD study team for their help and guidance in accessing the data. We are also grateful to James Groves, Rebecca Street, Tom Brown, Anjali Raghavan and Meera Sonara at the UCL Dementia Research Centre for their time recruiting participants to this study. We would like to thank Anna Joffe for her help in developing the Minder platform. We would like to acknowledge the Swallow group at University College London, who performed the DNA extractions.

## Author Contributions

MREC, JK-R and HM-S wrote the first draft of the manuscript and contributed to the design of the study. DJS and JMS planned the study, obtained funding, and supervised the study. MREC coordinated participant recruitment and data collection. JK-R recruited participants and responded to troubleshooting queries. MREC, JK-R, and ES produced figures and tables. LA managed the study and contributed to study design. All authors critically revised the manuscript for intellectual content.

## Funding

InSleep46 is primarily funded by the National Institute for Health and Care Research (NIHR204286).

Data collection for Phase 1 and 2 of Insight 46 was primarily funded by grants from Alzheimer’s Research UK (ARUK-PG2014-1946, ARUK-PG2017-1946), the Medical Research Council Dementias Platform UK (CSUB19166), the Wolfson Foundation (PR/ylr/18575), and UK Medical Research Council (MC_UU_00019/1, MC_UU_00019/3). Phase 3 was principally funded by the Alzheimer’s Association (SG-666374-UK BIRTH COHORT). Additional Support for the project comes from Brain Research UK (UCC14191), British Heart Foundation (PG/17/90/33415), National Brain Appeal, UK Dementia Research Institute (which receives its funding from UK DRI Ltd funded by the UK Medical Research Council, and the UCL/H Biomedical Research Centre). AVID Radiopharmaceuticals (a wholly owned subsidiary of Eli Lilly) provided the [18F]florbetapir Aβ PET tracer in kind during phases 1 and 2. Life Molecular Imaging, part of Life Healthcare’s Alliance Medical, provides the [18F]florbetaben Aβ PET tracer in kind during phase 3. Neither AVID Radiopharmaceuticals nor Life Molecular Imaging had any part in the design of the study.

JK-R is supported by a Medical Research Council Clinical Research Training Fellowship (MR/Y009452/1).

JMS is a National Institute for Health Research (NIHR) Senior Investigator and acknowledges the support of the NIHR University College London Hospitals Biomedical Research Centre and the UCL Centre of Research Excellence, an initiative funded by British Heart Foundation (RE/24/130013). This work is supported by the UK Dementia Research Institute through UK DRI Ltd, principally funded by the Medical Research Council. He has grant funding from Alzheimer’s Research UK, Brain Research UK, Weston Brain Institute, Medical Research Council, British Heart Foundation, Wolfson Foundation, and Alzheimer’s Association. This work uses data provided by study participants or patients and collected through clinical studies or as part of their care and support.

### RMA is funded by the NIHR (NIHR 301677) and NIHR Three Schools

Dementia Programme (Reference: 207100, NIHR-SSCR-DP27, NIHR-SSCR-DP-CDA30) and supported by the NIHR Newcastle Biomedical Research Centre based at The Newcastle upon Tyne Hospital National Health Service (NHS) Foundation Trust; Newcastle University; and the Cumbria, Northumberland and Tyne and Wear (CNTW) NHS Foundation Trust.

