## Supplementary material for "InSleep46: Deployment of a remote monitoring device for the detection and monitoring of dementia risk in older adult populations: a feasibility study"

**Supplementary Figure 1: Technological requirements for participation in InSleep46.**

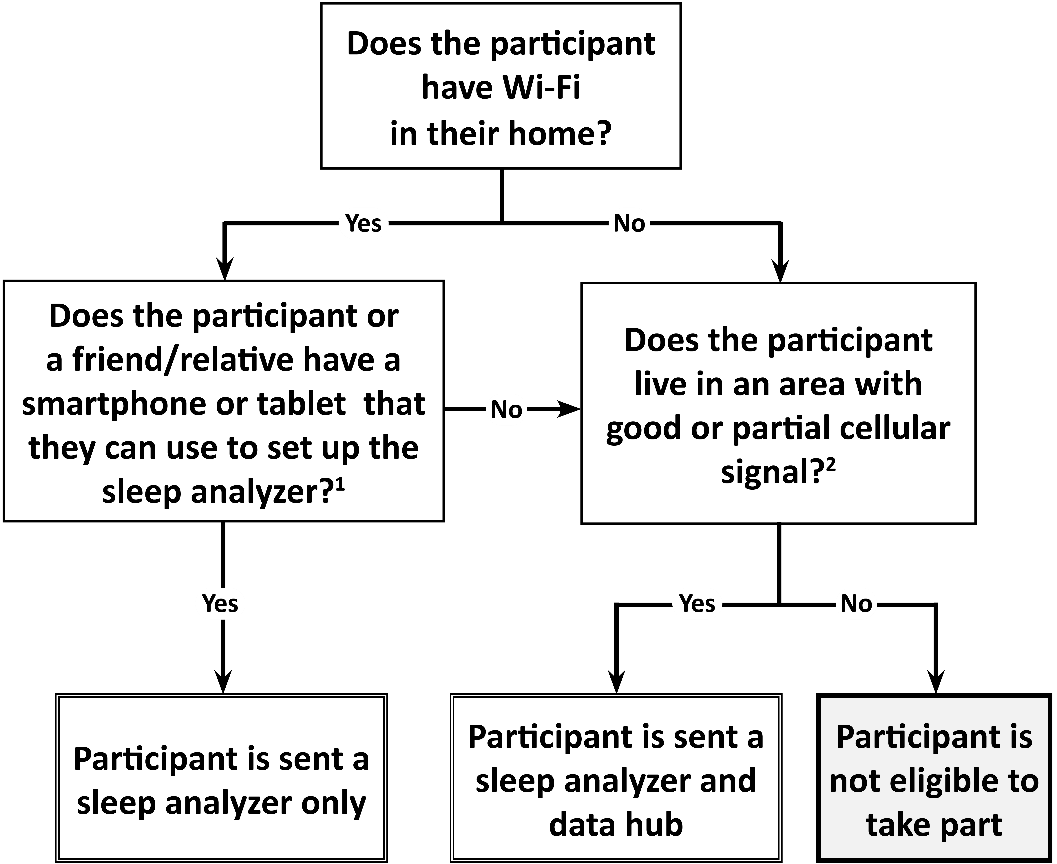
Flow chart showing technological requirements for participation in InSleep46. A smartphone/tablet^1^ is only required for initial device set up. Cellular signal^2^ was checked using the Ofcom mobile coverage checker: [www.ofcom.org.uk/mobile-coverage-checker](http://www.ofcom.org.uk/mobile-coverage-checker).

**Supplementary Table 1: Sleep habits questionnaire; asked to all participant during InSleep46 recruitment**

| **Question** | **Answers** |
| --- | --- |
| Do you have a typical side of the bed you sleep on? (when laying on back); | No; right; left; middle; diagonally; other. |
| What is the size of your bed? | Single; small double/queen; double; king; super king; Other |
| Do you have a typical sleeping position? | On back; on side; on front; varies; other. |
| How often do you have a bed partner (if at all)? | Never; Occasionally; Several nights per week; Every night. |
| How many nights per week do you sleep in your regular bed? | Occasionally; Several nights per week; Every night. |
| Does a pet sleep on your bed? | No; yes dog; yes cat; yes other. |
| Do you use an air conditioner to cool your home in the summer? | No; yes. |
| Do you heat your home in the winter? | No; yes. |
| Do you take any medications to help you sleep? | No; occasionally, when needed; every night |
| One hears about 'morning' and 'evening' types of people. The next questions relate to how you think of yourself in terms of being a 'morning' vs 'evening' person now, when you were in your 30s, and when you were in your 50s.   - Which one of these types do you consider yourself to be now? - Which one of these types do you consider yourself to be in your 30s? - Which one of these types do you consider yourself to be in your 50s? | For each of these questions, answers are: "Definitely a morning person",  "More a morning than an evening person", "More an evening person than a morning person  "Definitely an evening person"  "Do not know" (an answer is encouraged) |

**Supplementary Table 2: Troubleshooting issues and solutions.**

| Category | Examples of problem | Possible recommendations/solutions |
| --- | --- | --- |
| Technical issue – if using cellular connection | Loss of signal/no connection, cause unclear | 1. Confirm that both WSA and cellular hub are turned on 2. *If* light on cellular hub is flashing blue, the cellular connection is not established:    a. Confirm cellular coverage at participant’s home (<https://www.ofcom.org.uk/phones-and-broadband/coverage-and-speeds/ofcom-checker>).    b. Move cellular hub as close to the window as possible, where signal is generally stronger.    c. If light continues to flash blue, convert to Wi-Fi connection where available, this requires a reset of the WSA after attempt to pair with data hub (<https://mindermeetingplace.com/sleep-sensors/newwifi/>)   *If* light on cellular hub is green, it is connected to cellular network, the issue is likely related to the WSA or the connection to it:   a. Ensure that the cellular hub is in the same room as the WSA. b. If problem persists, reset the WSA . c. If problem persists, connect to Wi-Fi where available. |
| Technical issue – if using Wi-Fi | Loss of signal/no connection, cause unclear  Specific issues  Error due to attempted connection to 5 GHz band Wi-Fi.  Withings Connect Assistant app does not work with device.  WSA will not pair with phone/tablet.  Participant unable to download apps or similar technical limitations | 1. Check that the device has not accidentally been disconnected from power. 2. Confirm whether the participant has changed their router, necessitating re-pairing the WSA ((<https://mindermeetingplace.com/sleep-sensors/newwifi/>) 3. If neither of these is the cause, reset the WSA, and re-pair to Wi-Fi. 4. If still unsuccessful, trial cellular connection where available (<https://www.ofcom.org.uk/phones-and-broadband/coverage-and-speeds/ofcom-checker>).  - Guide participant to use the 2.4 GHz option, where available. Where this is not possible, support adding 2.4 GHz band, or separate the bands, where 2.4 GHz and 5 GHz are combined, if the router will allow this. - Attempt on an alternative device. Withings were informed of this issue and updated their app to fit wider range of devices  1. Ensure Bluetooth is switched on, and permission granted for Connect Assistant app to use it. 2. Try with alternative device. 1. Explain process (<https://mindermeetingplace.com/sleep-sensors/user-guide-wifi-connection/>) 2. Ask if a friend or relative can set up the device. Note that their smart device is only required for the setup. 3. If still unsuccessful, trial cellular connection where available (<https://www.ofcom.org.uk/phones-and-broadband/coverage-and-speeds/ofcom-checker>). |
| Technical issue - general | Anomalous/incomplete data received, such as intermittent absent when the participant confirms they were in bed. | - Check that the WSA has been placed correctly under the mattress in the correct position and is not placed directly on top of a slatted bed frame. |
| Participant medical issue | Participant had operation so unable to lift mattress to install WSA. Participant had fractured hip so sleeping in different bed  Participant had chest infection so hadn’t set up yet | 1. If appropriate, suggest that family member/friend can assist with set-up. 2. Plan follow up call at appropriate time interval |
| Social issue | Family member unwell/  holiday/awaiting assistance from family member | 1. Where appropriate, encourage set up and offer to talk through process. 2. Plan follow up call at appropriate time interval. |

**Supplementary Table 3: Predictors for InSleep46 recruitment and WSA connection; adjusted for sex, education attainment, childhood and adult SEP.**

Results of logistic regression analyses examining factors predictive of InSleep46 recruitment and Withings Sleep Analyzer (WSA) connection, adjusted for sex, education attainment, child and adult SEP. Analyses of sex, education attainment, child and adult socioeconomic position (SEP) were adjusted for the three remaining variables. OR, odds ratio; CI, confidence interval*,* PACC; Preclinical Alzheimer Cognitive Composite.

| **Variable** | **Recruited max n = 356** | | | **Connected max n = 263** | | |
| --- | --- | --- | --- | --- | --- | --- |
|  | **OR** | **p** | **95% CI** | **OR** | **p** | **95% CI** |
| **Sex** |  |  |  |  |  |  |
| Male | Reference |  |  | Reference |  |  |
| Female | 1.15 | 0.6 | 0.69, 1.90 | 0.44 | 0.1 | 0.16, 1.24 |
| **Childhood SEP** |  |  |  |  |  |  |
| Manual | Reference |  |  | Reference |  |  |
| Non-manual | 0.94 | 0.8 | 0.55, 1.62 | 1.26 | 0.7 | 0.45, 3.53 |
| **Educational attainment** |  |  |  |  |  |  |
| None | Reference |  |  | Reference |  |  |
| Up to O-level or equivalent (age 16) | 1.25 | 0.6 | 0.59, 2.66 | 0.75 | 0.7 | 0.14, 4.04 |
| A-level and above (age 16+) | 1.70 | 0.2 | 0.79, 3.66 | 0.71 | 0.7 | 0.13, 3.84 |
| **Adult SEP*** |  |  |  |  |  |  |
| Manual | Reference |  |  | Reference |  |  |
| Non-manual | 0.78 | 0.5 | 0.38, 1.60 | 0.77 | 0.7 | 0.16, 3.70 |
| **Childhood cognitive score** |  |  |  |  |  |  |
| Bottom 10% | Reference |  |  | Reference |  |  |
| Middle 80% | 1.26 | 0.6 | 0.54, 2.94 | 0.69 | 0.7 | 0.08, 6.08 |
| Top 10% | 1.07 | 0.9 | 0.34, 3.36 | 0.60 | 0.7 | 0.04, 8.48 |
| **Adult cognitive score, PACC** |  |  |  |  |  |  |
| Bottom 10% | Reference |  |  | Reference |  |  |
| Middle 80% | 1.97 | 0.2 | 0.78, 4.97 | 1.39 | 0.9 | 0.14, 14.02 |
| Top 10% | 4.65 | *0.05* | 0.10, 21.66 | 2.47 | 0.6 | 0.11, 56.90 |
| **Smoking exposure** |  |  |  |  |  |  |
| Never smoked | Reference |  |  | Reference |  |  |
| Ex or current smoker | 0.69 | 0.2 | 0.41, 1.17 | 0.67 | 0.5 | 0.24, 1.87 |
| **Alcohol use at age 76** |  |  |  |  |  |  |
| Never | Reference |  |  | Reference |  |  |
| Up to once per week | 0.75 | 0.6 | 0.29, 1.95 | 0.58 | 0.6 | 0.07, 5.25 |
| 2-3 times per week | 1.07 | 0.9 | 0.39, 2.93 | 0.54 | 0.6 | 0.06, 5.00 |
| 4+ times per week | 0.83 | 0.7 | 0.32, 2.18 | 0.65 | 0.7 | 0.07, 6.08 |
| **Self-rated health at age 76-77** |  |  |  |  |  |  |
| Poor or Fair | Reference |  |  | Reference |  |  |
| Good | 1.28 | 0.6 | 0.57, 2.84 | 2.63 | 0.2 | 0.53, 13.02 |
| Very good or excellent | 1.47 | 0.3 | 0.68, 3.18 | 1.62 | 0.5 | 0.39, 6.67 |
| ***APOE* status** |  |  |  |  |  |  |
| Negative | Reference |  |  | Reference |  |  |
| Positive (heterozygous or homozygous) | 0.75 | 0.3 | 0.44, 1.28 | 2.06 | 0.4 | 0.44, 9.65 |
| **Functional independence** |  |  |  |  |  |  |
| Independent | Reference |  |  | Reference |  |  |
| Dependent in ≥1 domain | 1.02 | 1.0 | 0.26, 4.04 | 0.20 | *0.08* | 0.03, 1.22 |
